# Safety and effectiveness of COVID-19 mRNA vaccination and risk factors for hospitalisation caused by the omicron variant in 0.8 million adolescents: A nationwide cohort study in Sweden

**DOI:** 10.1101/2022.10.19.22281286

**Authors:** Peter Nordström, Marcel Ballin, Anna Nordström

## Abstract

**Background:** Real-world evidence on the safety and effectiveness of COVID-19 vaccination against severe disease caused by the omicron variant among adolescents is sparse. In addition, evidence on risk factors for severe COVID-19 disease, and whether vaccination is similarly effective in such risk groups, is unclear.

**Methods and findings:** Nationwide registers were used to examine the safety and effectiveness of COVID-19 mRNA vaccination against COVID-19 hospitalisation, and risk factors for COVID-19 hospitalisation in adolescents. The safety analysis included all individuals in Sweden born between 2003-2009 (aged 11.3-19.2 years) given at least one dose of mRNA vaccine (N=645,355), and never vaccinated controls (N=199,022). Outcomes evaluated included all hospitalisations until 5 June 2022. The vaccine effectiveness (VE) against COVID-19 hospitalisation and associated risk factors was evaluated in adolescents given two doses of mRNA vaccine (N=501,945), as compared to never vaccinated controls (N=170,083), during an omicron predominant period (1 January 2022 to 5 June 2022). The safety analysis showed that COVID-19 mRNA vaccination was not associated with an increased risk of any serious adverse events resulting in hospitalisation. During follow-up, 1.69% of the vaccinated individuals were hospitalised compared to 1.71% of the controls (P=0.29). In the VE analysis, there were 21 cases (0.004%) of COVID-19 hospitalisation among 2-dose recipients and 26 cases (0.015%) among controls, resulting in an estimated VE of 75% (95% CI, 54-86, P<0.001). Strong risk factors for COVID-19 hospitalisation included previous infections (odds ratio [OR], 14.3, 95% CI, 7.7-26.6, P<0.001), and cerebral palsy/development disorders (OR, 12.0, 95% CI, 6.4-22.6, P<0.001), with similar estimates of VE in these subgroups as in the total cohort. The number needed to vaccinate with two doses to prevent one case of COVID-19 hospitalisation was 9,007 in the total cohort and 1,031 in those with previous infections or developmental disorders. None of the individuals hospitalised due to COVID-19 died within 30 days.

**Conclusions:** In this nationwide study, COVID-19 mRNA vaccination was not associated with an increased risk of any serious adverse event in adolescents. Two doses were associated with a lower risk of COVID-19 hospitalisation during the omicron predominant period, especially among those with certain predisposing conditions who should be prioritized for vaccination. However, COVID-19 hospitalisation among general adolescents was extremely rare, and additional doses in this population may not be warranted at this stage.

**Why was this study done:** *Evidence before this study:*

➢ There is limited evidence on the effectiveness of COVID-19 vaccination against severe outcomes during the omicron era among adolescents. In addition, there is lack of data on whether certain groups of adolescents are at greater risk of severe COVID-19 and should be prioritized in vaccination programs, and whether vaccination is equally effective in such risk groups.
➢ Regarding safety, some studies have indicated a link between COVID-19 mRNA vaccination and increased risk of myocarditis and pericarditis in young men, although the data appear inconsistent.

*What did the researchers do and find?:*

➢ Using Swedish nationwide health registers, a cohort of 844,377 adolescents were followed until 5 June 2022 to evaluate the safety and effectiveness of COVID-19 mRNA vaccination against COVID-19 hospitalisation, and risk factors for COVID-19 hospitalisation.
➢ COVID-19 mRNA vaccination in adolescents was not associated with an increased risk of any diagnoses resulting in hospitalisation. In contrast, two doses of vaccine had an associated 75% effectiveness against COVID-19 hospitalisation. However, only about 6 individuals in 100,000 were hospitalised due to COVID-19 during follow-up.
➢ There were certain strong risk factors for COVID-19 hospitalisation, such as previous infections and different development disorders, which increased the risk of COVID-19 hospitalisation more than tenfold. Vaccine effectiveness among these individuals was similar as in the rest of the cohort.

*What do these findings mean?:*

➢ Although COVID-19 mRNA vaccination appears safe and associated with reduced risk of COVID-19 hospitalisation, the risk of severe COVID-19 seems to be extremely low in general adolescents. Therefore, administration of additional doses to the general population of adolescents may not be warranted at this stage of the pandemic. In contrast, individuals with a high risk for severe COVID-19 should be prioritised for vaccination.

## Introduction

The omicron variant of SARS-CoV-2 led to a surge in cases among young people during 2022 when several countries lifted or alleviated their restrictions.[1] Although clinical trials showed that the BNT162b2 and mRNA-1273 vaccines had acceptable safety profiles in adolescents and reduced the risk of infection in the short-term,[2, 3] these trials were conducted before the emergence of omicron. Limited sample sizes also hindered the evaluation of the level of protection against severe COVID-19, such as hospitalisation, which could be estimated using large-scale observational studies. These studies also offer the possibility of investigating any potential links between vaccination and rare serious adverse events, such as myocarditis, for which the data appear inconsistent.[4, 5]

Currently, there is limited data on vaccine effectiveness (VE) against severe COVID-19 caused by the omicron variant among adolescents. Two case-control studies found that two doses of the BNT162b2 mRNA vaccine had about 80% VE against COVID-19 hospitalisation or death in adolescents.[6] [7] However, given their study design, it is difficult to determine how common severe disease is during the omicron era. In addition, little is known about whether certain groups of adolescents should be prioritized for vaccination because of a higher risk of severe COVID-19, and whether vaccination has a similarly protective effect in such risk groups. Moreover, although a third dose, also known as a booster dose, may increase protection against symptomatic omicron infection in adolescents,[8, 9] the risk of severe COVID-19 after a third dose relative to after the second dose is unclear.

Therefore, the present study leveraged Swedish nationwide registers to evaluate, among adolescents, 1) the risk of serious adverse events following COVID-19 mRNA vaccination, and 2) the effectiveness of COVID-19 mRNA vaccination against hospitalisation due to COVID-19 and risk factors for COVID-19 hospitalisation during an omicron predominant period.

## Methods

### Study design and cohorts

This nationwide, retrospective cohort study was approved by the Swedish Ethical Review Authority (number 00094/2021), who waived the requirement of obtaining informed consent given the retrospective study design.

The cohort considered for inclusion was compiled by Statistics Sweden (www.SCB.se); the government agency in charge of nationwide statics in different areas and covering the total population of Sweden. All individuals born 2003-2009 who were given at least one dose of COVID-19 vaccine or had a documented SARS-CoV-2 infection until March 2022 were considered for inclusion (N=692,333). To these individuals, Statistics Sweden matched one control individual on birth year, sex, and municipality. Controls could not have a documented SARS-CoV-2 infection at the date of documented infection in the corresponding infected individual, and they could not have received a first dose of vaccine at the date when the corresponding vaccinated individual had received two doses of vaccine. The cohort was updated with vaccination data and SARS-CoV-2 infections until 2 June 2022. Because one control could be matched to several vaccinated individuals, the total eligible cohort consisted of 844,511 individuals, of whom 645,489 had been vaccinated with at least one dose, 600,846 had been vaccinated with two doses, and 60,475 had been vaccinated with at least three doses (Figure 1). After excluding a few individuals vaccinated with a vector-based vaccine (N=134), a total of 844,377 individuals remained and were included in the present study for two sets of analyses. In a first set of analyses, named the safety analysis, all diagnoses set during hospitalisation were evaluated in all individuals given at least one dose of vaccine during follow-up (N=645,355) as compared to individuals never vaccinated during follow-up (N=199,022). In a second set of analyses, VE against COVID-19 hospitalisation was evaluated by comparing individuals given at least two doses of vaccine (N=501,945) to individuals never vaccinated during follow-up (N=170,083), excluding all individuals with a previous documented SARS-CoV-2 infection (Figure 1). Data on individuals vaccinated against COVID-19 and data on documented SARS-CoV-2 infections were collected from the Swedish Vaccination Register and the SmiNet register, respectively. Both these registers are managed by the Public Health Agency of Sweden and all healthcare providers in Sweden were obliged to report to these registers according to law.[10, 11]

**Figure 1.**
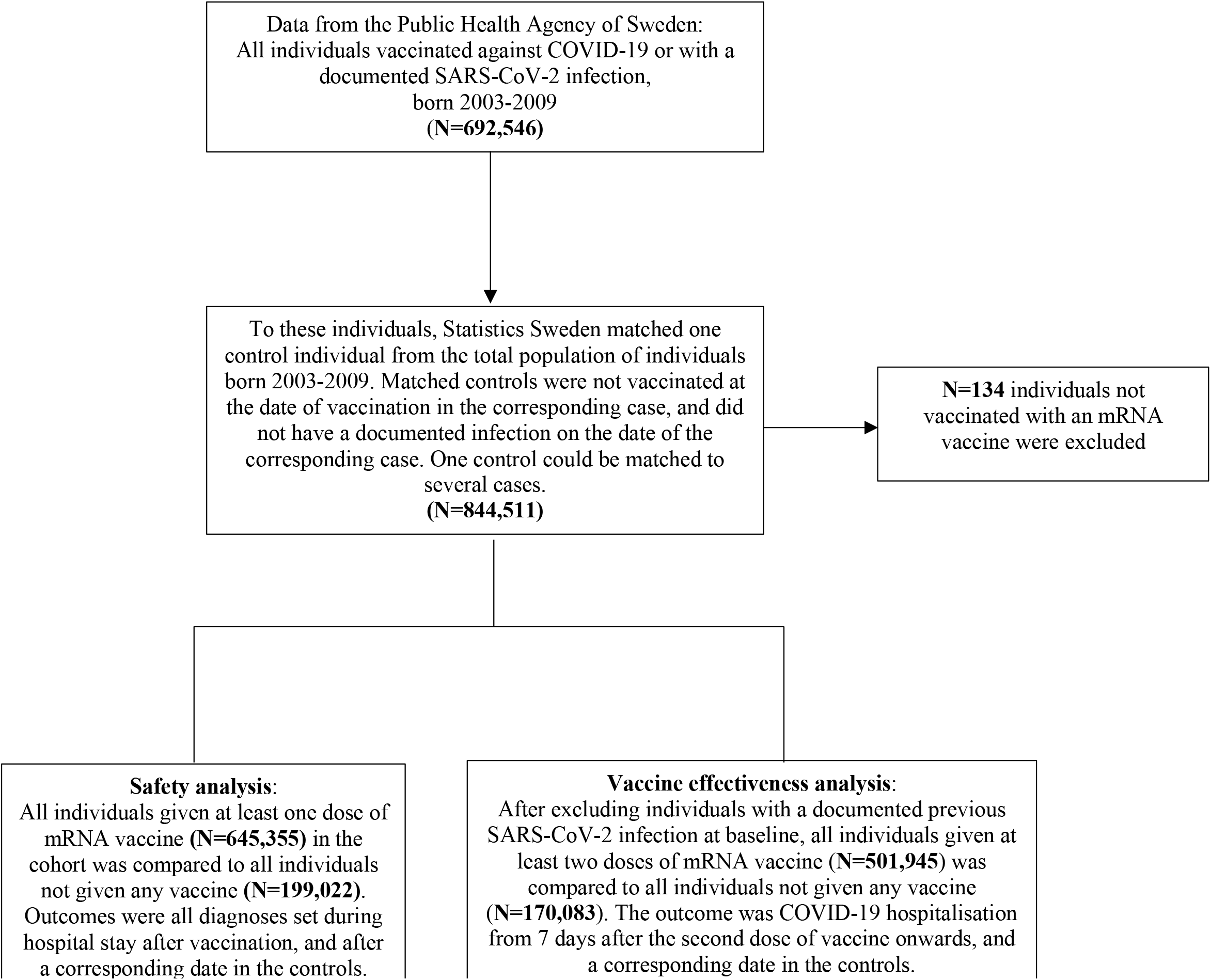
Selection of study participants.

### Outcomes

In the safety analysis, based on all diagnoses set in the cohort during the hospital stay until 5 June 2022, results are presented for 30 different diagnoses. These diagnoses were selected based on their incidence and general interest given previous reports of links between certain diagnoses and COVID-19 mRNA vaccination.[4, 12, 13] Only the first diagnosis was evaluated for each individual, and individuals with this diagnosis at baseline were therefore excluded from the prospective analyses.

In the VE analysis, the primary outcome was a main diagnosis of COVID-19 (thus, “due to” COVID-19 rather than “with” COVID-19), set during inpatient hospital stay from 1 January 2022 until 5 June 2022. The secondary outcome was documented SARS-CoV-2 infection of any severity from the SmiNet register, from 1 January 2022 until 28 February 2022, as per when the guidelines for testing in Sweden had changed. For these two outcomes, only cases that occurred 7 days after the second dose of vaccine and onwards were counted to ensure the full effect of vaccination. The start of 1 January was selected on the basis that the omicron variant was first documented in Sweden on 29 November 2021[14] and by early January 2022 it represented >90% of sequenced cases (Supplementary Table 1). Data on all diagnoses used in this study were obtained from the National Patient Register[15] using 10^th^ revision International Classification of Disease codes, starting from 1 January 2016 earliest until 5 June 2022 latest. The definition and diagnostic codes for all diagnoses considered in the safety analysis and for the analysis of risk factors for COVID-19 hospitalisation are shown in Table 1. The positive predictive value for diagnoses set within the National Patient Register differs, but has generally been found to be between 85-95%, although sensitivity is often lower.[15] Finally, data on death due to COVID-19 or influenza, defined as death within 30 days after the diagnosis, were obtained from the National Cause of Death Register.[16] The present study was approved by the Swedish Ethical Review Authority.

**Table 1.**
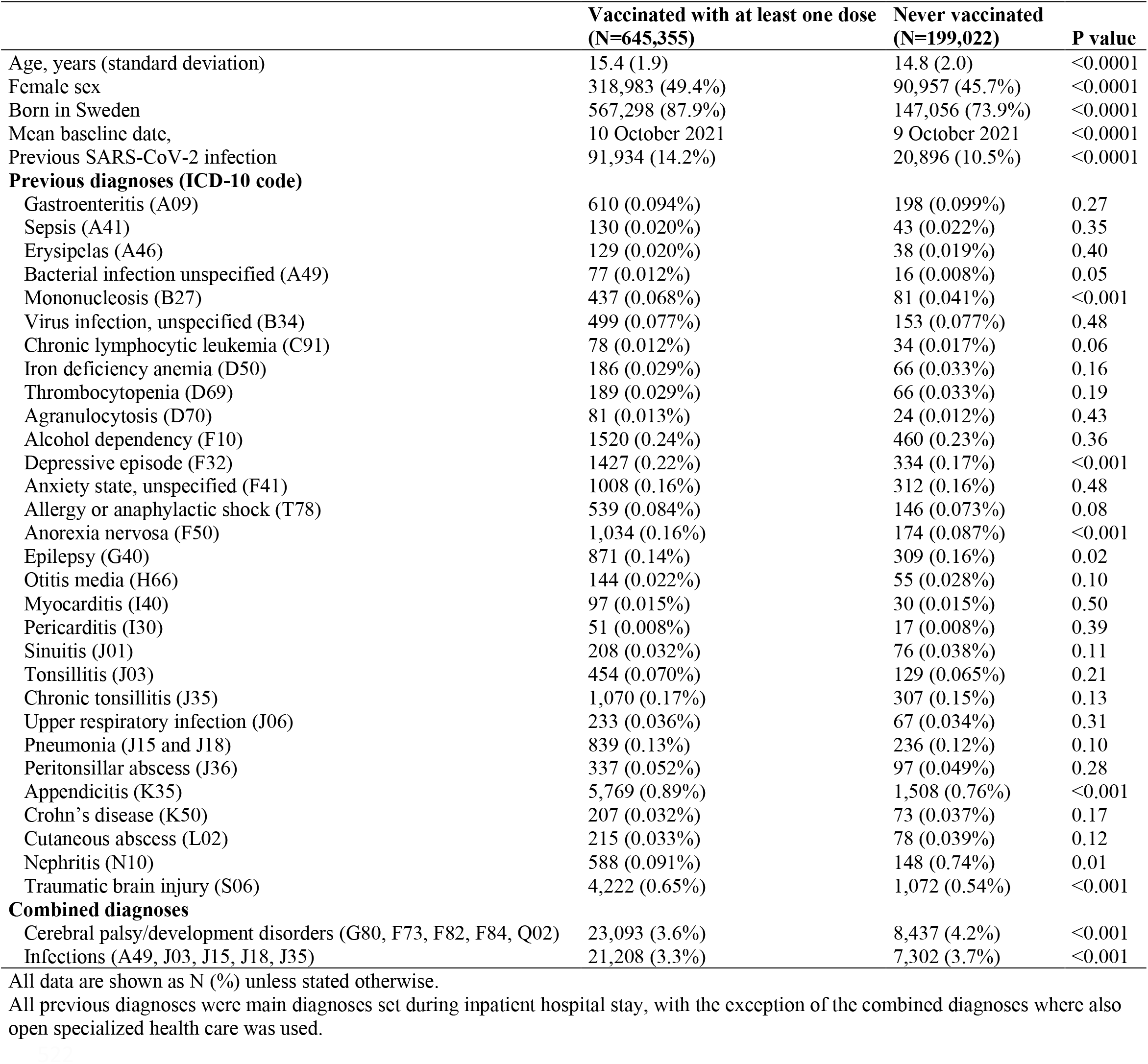
Baseline characteristics of the individuals at date of the first dose of vaccine and in never vaccinated individuals.

### Statistical analysis

In the safety analysis, diagnoses were evaluated before and after the first dose of vaccine in 645,355 individuals given at least one dose of vaccine and in 199,022 controls who were unvaccinated during the follow-up time. The baseline date in vaccinated individuals was the date of vaccination with the first dose. In the controls, a baseline date was randomly assigned based on the mean baseline date and standard deviation among the vaccinated individuals (10 October 2021 ± 54 days). Student’s t-tests and chi-square tests were used to compare the prevalence of different variables at baseline. To estimate hazard ratios (HRs) for the 30 different diagnoses during follow-up, Cox regression models were used. Individuals were censored on the date of the diagnosis of interest, death, or end of follow-up (5 June 2022), whichever came first.

In the VE analysis, logistic regression was used to estimate odds ratios (OR) for the primary outcome of COVID-19 hospitalisation (from 1 January 2022 until 5 June 2022), and for the secondary outcome of a SARS-CoV-2 infection (from 1 January 2022 until 28 February 2022). The ORs obtained were used to estimate VE as 1 minus the OR x 100. In all analyses, the first model was unadjusted and the second model was adjusted for baseline date, age, sex, and whether the individual was born in Sweden or not. Data underlying these covariates were retrieved from Statistics Sweden.[17] To investigate whether VE differed by the covariates, interaction analyses were performed using product terms created by multiplying the variable coding for vaccination status at baseline (vaccinated/unvaccinated) by each respective covariate, which was added to the logistic regression model. Given that the interaction term was statistically significant (P<0·05) for the baseline date, VE was also estimated in subgroups according to this covariate. The number needed to vaccinate (NNV) with two doses to prevent one case of COVID-19 hospitalisation during follow-up was estimated as the inverse of the absolute risk difference between the groups (vaccinated/unvaccinated). The NNV was estimated both in the total cohort and by subgroups.

Finally, a sensitivity analysis using a negative control outcome was conducted to explore the potential risk of bias due to unmeasured confounding.[18] Here, a logistic regression model was performed, comparing individuals given at least two doses of COVID-19 mRNA vaccine compared to never vaccinated individuals concerning the outcome of hospitalisation due to influenza from 1 January 2022 until 5 June 2022. All analyses were performed in SPSS v29·0 for Mac (IBM Corp, Armonk, NY, USA), and Stata v16·1 for Mac (Statcorp, College Station, Texas, USA). A two-sided P-value <0·05 or Ors/HRs with 95% CIs not crossing one were considered statistically significant.

## Results

The total cohort comprised 844,377 adolescents born 2003-2009 (age 11.3-19.2 years), of whom 645,355 received at least one dose of COVID-19 mRNA vaccine and 199,022 never vaccinated individuals (controls). Almost 90% of the vaccinated individuals received BNT162b2 as a first dose, while the remaining received mRNA-1273. Baseline characteristics are shown in Table 1. Individuals that were never vaccinated were slightly younger, more often of male sex and born outside of Sweden, and were less often diagnosed with a previous SARS-CoV-2 infection (P<0.001 for all). Concerning other diagnoses at baseline, differences between the groups were marginal (Table 1).

### Serious adverse events after a first dose of vaccine

In the safety analysis (N=844,377), there were a total of 19,660 hospitalisations among 14,320 individuals during follow-up. In vaccinated individuals, 1.69% (N=10,918) were hospitalised at least once, compared to 1.71% (N=3,402) in those never vaccinated (P=0.29 for difference). There were marginal differences between the two groups in the 30 diagnoses presented (Table 2), although statistically significant associations in favour of vaccination were observed with respect to the risk of sepsis, thrombocytopenia, alcohol dependency, and Crohn’s disease (P<0.05 for all). None of the other associations were statistically significant.

**Table 2.** Hazard ratios for 30 different diagnoses during follow-up in individuals vaccinated with at least one dose compared to never vaccinated individuals.

|  | Vaccinated with at least one dose (N=645,355) | Never vaccinated (N=199,022) |  |  |
| --- | --- | --- | --- | --- |
| Diagnosis (ICD-10 code) | Number of cases (incidence rate) | Number of cases (incidence rate) | HR (95% CI) | P value |
| Gastroenteritis (A09) | 58 (0.38) | 20 (0.43) | 1.02 (0.59-1.77) | 0.94 |
| Sepsis (A41) | 5 (0.03) | 7 (0.15) | 0.18 (0.06-0.59) | 0.004 |
| Erysipelas (A46) | 8 (0.05) | 2 (0.04) | 1.04 (0.19-5.67) | 0.97 |
| Bacterial infection unspecified (A49) | 11 (0.07) | 3 (0.06) | 0.87 (0.24-3.21) | 0.84 |
| Mononucleosis (B27) | 115 (0.76) | 26 (0.55) | 0.82 (0.53-1.26) | 0.36 |
| Virus infection, unspecified (B34) | 42 (0.28) | 7 (0.15) | 1.65 (0.73-3.73) | 0.23 |
| Chronic lymphocytic leukemia (C91) | 4 (0.03) | 4 (0.09) | 0.36 (0.08-1.50) | 0.16 |
| Iron deficiency anemia (D50) | 38 (0.25) | 14 (0.30) | 0.94 (0.50-1.76) | 0.84 |
| Thrombocytopenia (D69) | 8 (0.05) | 10 (0.21) | 0.22 (0.08-0.60) | 0.003 |
| Agranulocytosis (D70) | 1 (0.007) | 1 (0.02) | - | - |
| Alcohol dependency (F10) | 409 (2.69) | 124 (2.65) | 0.79 (0.64-0.97) | 0.02 |
| Depressive episode (F32) | 363 (2.39) | 103 (2.19) | 0.92 (0.73-1.15) | 0.46 |
| Anxiety state, unspecified (F41) | 341 (2.25) | 75 (1.60) | 0.96 (0.74-1.24) | 0.76 |
| Allergy or anaphylactic shock (T78) | 64 (0.42) | 23 (0.49) | 0.74 (0.45-1.21) | 0.23 |
| Anorexia nervosa (F50) | 197 (1.30) | 48 (1.02) | 1.12 (0.81-1.55) | 0.50 |
| Epilepsy (G40) | 82 (0.54) | 34 (0.73) | 0.89 (0.57-1.40) | 0.62 |
| Otitis media (H66) | 7 (0.05) | 4 (0.09) | 0.61 (0.16-2.33) | 0.47 |
| Myocarditis (I40) | 86 (0.57) | 19 (0.41) | 1.05 (0.63-1.74) | 0.86 |
| Pericarditis (I30) | 21 (0.14) | 3 (0.06) | 0.92 (0.27-3.17) | 0.89 |
| Sinuitis (J01) | 15 (0.10) | 10 (0.21) | 0.68 (0.27-1.69) | 0.41 |
| Tonsillitis (J03) | 90 (0.59) | 11 (0.23) | 1.58 (0.83-3.00) | 0.16 |
| Chronic tonsillitis (J35) | 94 (0.59) | 19 (0.41) | 1.17 (0.70-0.94) | 0.56 |
| Upper respiratory infection (J06) | 24 (0.16) | 5 (0.11) | 1.64 (0.57-4.73) | 0.36 |
| Pneumonia (J15 and J18) | 68 (0.45) | 17 (0.36) | 1.47 (0.83-2.60) | 0.19 |
| Peritonsillar abscess (J36) | 52 (0.34) | 17 (0.36) | 0.58 (0.33-1.03) | 0.06 |
| Appendicitis (K35) | 665 (4.38) | 176 (3.76) | 1.09 (0.92-1.29) | 0.34 |
| Crohn's disease (K50) | 24 (0.16) | 14 (0.30) | 0.48 (0.24-0.97) | 0.04 |
| Cutaneous abscess (L02) | 10 (0.07) | 5 (0.11) | 0.63 (0.21-1.90) | 0.41 |
| Nephritis (N10) | 105 (0.69) | 19 (0.41) | 1.21 (0.73-1.99) | 0.47 |
| Traumatic brain injury (S06) | 307 (2.02) | 86 (1.84) | 1.13 (0.88-1.45) | 0.35 |

CI = confidence interval, HR = hazard ratio.
Incidence rates are presented as cases per one million person-years. HRs were adjusted for age, sex, baseline date, and whether the individual was born in Sweden.

### Vaccine effectiveness against COVID-19 hospitalisation

In the analysis of VE against COVID-19 hospitalisation, 501,945 individuals vaccinated with two doses and 170,083 never vaccinated controls were included. Between 1 January 2022 and 5 June 2022, a total of 47 individuals (6 per 100,000) were hospitalised due to COVID-19. Of these, 21 cases were among those vaccinated with two doses (0.004%), and 26 among unvaccinated (0.015%), resulting in a VE of 75% (95% CI, 54-86, P<0.001) (Table 3). The NNV with two doses to prevent one case of COVID-19 hospitalisation was 9,007. For those with a second dose of vaccine earlier than 15 November, the VE was 68% (95% CI, 25-86, P=0.009), compared to 86% (95% CI, 63-95, P<0.001) for those with a second dose of vaccine 15 November 2021 and later (P=0.03 for interaction). When comparing individuals vaccinated with three doses (N=41,225) compared to those vaccinated with two doses (N=413,544), only 12 individuals were hospitalised due to COVID-19 during follow-up (VE, 13%, 95% CI, -354-84, P=0.86). There were no deaths within 30 days of hospitalisation among the 238 individuals hospitalised due to COVID-19 in the total cohort (N=844,377) since the beginning of the pandemic in January 2020.

**Table 3.** Vaccine effectiveness against COVID-19 hospitalisation from 7 days onwards after a second dose of vaccine as compared to never vaccinated individuals from 1 January 2022 until 5 June 2022, and by time since vaccination and subgroups.

|  | Vaccinated<br>with two doses | Never<br>vaccinated | Unadjusted<br>vaccine<br>effectiveness<br>(95% CI) | Adjusted<br>vaccine<br>effectiveness<br>(95% CI) |
| --- | --- | --- | --- | --- |
|  | Number of<br>cases (%) | Number of<br>cases (%) |  |  |
| **Total cohort (N=672,028)<sup>a</sup>** | 21 (0.004%) | 26 (0.015%) | 73 (51-85) | 75 (54-86) |
| **Subgroups** |  |  |  |  |
| Baseline date < 15 Nov 2021 (N=293,643)<sup>b</sup> | 15 (0.007%) | 14 (0.017%) | 58 (13-80) | 68 (25-86) |
| Baseline date > 14 Nov 2021 (N=378,385)<sup>c</sup> | 6 (0.002%) | 12 (0.014%) | 85 (60-94) | 86 (63-95) |
| Previously diagnosed with infections (N=22,125)<sup>d</sup> | 4 (0.025%) | 11 (0.182%) | 86 (57-96) | 88 (57-96) |
| Previously diagnosed with development disorders(N=25,962)<sup>e</sup> | 6 (0.032%) | 9 (0.121%) | 73 (15-90) | 72 (18-90) |

CI = confidence interval.
Adjusted models were adjusted for age, baseline date, sex and whether the individual was born in Sweden.
<sup>a</sup>Of which 501,945 were vaccinated and 170,083 never vaccinated.
<sup>b</sup>Of which 210,747 were vaccinated and 82,896 never vaccinated.
<sup>c</sup>Of which 291,198 were vaccinated and 87,187 never vaccinated.
<sup>d</sup>Of which 16,083 were vaccinated and 6,042 never vaccinated.
<sup>e</sup>Of which 18,553 were vaccinated and 7,409 never vaccinated.

### Risk factors for COVID-19 hospitalisation and vaccine effectiveness by subgroups

Of the 47 individuals hospitalised due to COVID-19 in the VE analysis, 28 (60%) had previously been hospitalised for another condition, compared to 66,756 (10%) of the individuals in the rest of the cohort. Among the individuals hospitalised due to COVID-19, 15 (32%) had previously been diagnosed with an infection, compared to 22,110 (3.3%) in the rest of the cohort, equal to an adjusted OR for COVID-19 hospitalisation of 14.3 (95% CI, 7.7-26.6, P<0.001), after adjustment for age, sex, baseline date, vaccination status, and whether the individual was born in Sweden. The VE in this risk group was similar (VE, 88%, 95% CI, 57-96, P<0.001), as in the total cohort, but with a slightly lower vaccination coverage (72.7% vs. 74.7%). In addition, 15 (32%) of the individuals hospitalised due to COVID-19 had previously been diagnosed with cerebral palsy and/or different development disorders, compared to 25,947 individuals (3.9%) in the rest of the cohort, resulting in an adjusted OR for COVID-19 hospitalisation of 12.0 (95% CI, 6.4-22.6, P<0.001). Again, VE in this subgroup was similar as in the total cohort (VE, 72%, 95% CI, 18-90, P=0.02), but with a slightly lower vaccination coverage (71.5% vs 74.8%). The NNV with two doses to prevent one case of COVID-19 hospitalisation in the subgroup of individuals previously diagnosed with infections or development disorders (N=46,783) was 1,031.

### Vaccine effectiveness against SARS-CoV-2 infection

The VE against SARS-CoV-2 infection of any severity was estimated among 488,441 individuals vaccinated with two doses compared to 165,626 never vaccinated controls. Between 1 January 2022 and 28 February 2022, there were 72,631 cases of confirmed SARS-CoV-2 infection. The VE varied by time since the last dose (P<0.001), with no VE in the total cohort (VE, -2%, 95% CI, -4-0, P=0.02) (Table 4). There was a marginal VE among individuals with a second dose no earlier than 1 September 2021 (VE, 4%, 95% CI, 2-6, P<0.001), a low VE for individuals with a second dose no earlier than 1 November 2021 (VE, 21%, 95% CI, 19-23, P<0.001), and a moderate VE for individuals with a second dose no earlier than 1 January 2022 (VE, 47%, 95% CI, 43-51, P<0.001).

**Table 4.** Vaccine effectiveness against SARS-CoV-2 infection of any severity from 7 days onwards after a second dose of vaccine as compared to never vaccinated individuals from 1 January 2022 until 28 February 2022, and by time since vaccination.

|  | Vaccinated<br>with two doses | Never vaccinated | Unadjusted<br>vaccine<br>effectiveness<br>(95% CI) | Adjusted<br>vaccine<br>effectiveness<br>(95% CI) |
| --- | --- | --- | --- | --- |
|  | Number of<br>cases (%) | Number of<br>cases (%) |  |  |
| **Total cohort (N=654,067)<sup>a</sup>** | 55,101 (11.3%) | 17,530 (10.6%) | -7 (-9, 6) | -2 (-4, 0) |
| **Subgroups** |  |  |  |  |
| ≥ 1 September 2021 (N=622,296)<sup>b</sup> | 51,497 (11.0%) | 16,037 (10.5%) | -5 (-7, -3) | 4 (2, 6) |
| ≥ 1 October 2021 (N=487,348)<sup>c</sup> | 32,613 (9.2%) | 13,655 (10.3%) | 11 (9, 13) | 10 (8, 12) |
| ≥ 1 November 2021 (N=409,833)<sup>d</sup> | 27,213 (8.8%) | 9,743 (9.7%) | 11 (9, 13) | 21 (19, 23) |
| ≥ 1 December 2021 (N=245,914)<sup>e</sup> | 12,547 (6.8%) | 5,288 (8.5%) | 21 (18, 23) | 36 (34, 23) |
| ≥ 1 January 2022 (N=107,773)<sup>f</sup> | 2,087 (2.6%) | 1,279 (4.5%) | 42 (38, 46) | 47 (43, 51) |

CI = confidence interval. Adjusted models were adjusted for age, baseline date, sex and whether the individual was born in Sweden. <sup>a</sup>Of which 488,441 were vaccinated and 165,626 never vaccinated. <sup>b</sup>Of which 469,627 were vaccinated and 152,669 never vaccinated. <sup>c</sup>Of which 354,304 were vaccinated and 133,044 never vaccinated. <sup>d</sup>Of which 309,804 were vaccinated and 100,029 never vaccinated. <sup>e</sup>Of which 183,462 were vaccinated and 62,452 never vaccinated. <sup>f</sup>Of which 79,161 were vaccinated and 28,612 never vaccinated.

### Sensitivity analysis

The risk of hospitalisation due to influenza was evaluated in 600,021 individuals given at least two doses of vaccine compared to in 199,003 never vaccinated individuals. Between 1 January 2022 and 5 June 2022, a total of 48 individuals (6 per 100,000) were hospitalised due to influenza, with no detectable VE (VE, -6%, 95% CI, -51-119, P=0.88). Of the 130 individuals in the total cohort hospitalised for influenza since the start of the COVID-19 pandemic in January 2020, one individual died within 30 days of hospitalisation.

## Discussion

In this nationwide study of more than 0.8 million adolescents, COVID-19 mRNA vaccination was not associated with an increased risk of hospitalisation for any diagnosis, with an overall risk of hospitalisation which was similar in vaccinated and unvaccinated individuals. The results also showed that although vaccination was associated with lower risk of COVID-19 hospitalisation during an omicron predominant period, the absolute risk of being hospitalised was extremely low, except in subgroups of adolescents that had previously been diagnosed with infections, cerebral palsy or other development disorders.

Evidence on the safety and effectiveness of COVID-19 mRNA vaccination in adolescents during the omicron predominant period is limited. In this study, no association between vaccination and increased risk of hospitalisation in general, or for any specific diagnosis, was detected. This suggests that COVID-19 vaccination in adolescents is safe. These findings add to, and extend upon those from a nationwide study in Scotland, reporting no association between vaccination and increased risk of hospital stay for 17 different diagnoses among adolescents.[5] The present study also estimated that two doses of mRNA vaccine had about 75% effectiveness against hospitalisation due to COVID-19. Similar estimates were reported in two case-control studies of adolescents conducted in the US and Brazil during an omicron predominant period, where VE against COVID-19 hospitalisation or death was about 80%.[6] [7] However, given their design, these studies were unable to determine how common severe COVID-19 is. Therefore, the very low absolute risk of severe disease observed in the present cohort study based on a nationwide sample of adolescents is an important finding for decision making concerning the need for COVID-19 vaccination in adolescents. Overall, only 47 individuals, or 6 in 100,000, were hospitalised due to COVID-19 during a period of 5 months, and none of all adolescents hospitalised due to COVID-19 since the start of the pandemic died within 30 days of hospitalisation. Based on this very low risk of severe COVID-19 in the total cohort, the NNV with two doses to prevent one case during follow-up was about 9,000. Interestingly, the absolute risk of being hospitalised due to COVID-19 and influenza was similar, despite that the infection pressure from SARS-CoV-2 during follow-up was more than 100 times higher than that of influenza.[19]

Given the above, it is important to evaluate whether certain groups of adolescents are at higher risk of severe COVID-19, and if so, whether vaccination is associated with similar protection among these individuals. This study identified two different clusters of diagnoses which increased the risk of COVID-19 hospitalisation more than tenfold. The first included previous infections and the second included cerebral palsy and other developmental disorders. An encouraging finding was that the VE in these subgroups was similar to that in the total cohort, and consequently, the NNV to prevent one case of COVID-19 hospitalisation was about 1,000 individuals. It is therefore of concern that vaccination coverage in these risk groups was somewhat lower compared to in the total cohort. Although we are unable to determine the underlying causes for this observation, factors such as recurrent infections interfering with the administration of the vaccine, and fear of adverse events, may have contributed. Taken together, these findings suggest that vaccination of adolescents during the omicron era should primarily be targeting risk groups, such as those with previous infections and development disorders.

Concerning the outcome of SARS-CoV-2 infection of any severity, the results indicated that VE from primary vaccination wanes within a few months, similar to observations made in a few other countries.[7-9] However, there is a lack of data exploring whether booster doses reduce the risk of severe COVID-19 as compared to primary vaccination.[20] The results for this comparison in the present study showed that COVID-19 hospitalisations during follow-up were once again extremely rare, implying that administration of booster doses to the general population of adolescents may not be warranted at the current stage of the pandemic. Based on these findings, it is noteworthy that the US and several countries in Europe are recommending booster doses to adolescents.[21, 22]

The present study has limitations that should be considered. Because of the observational design, conclusions based on the associations found should be made with caution. For example, despite that VE changed marginally before and after adjustment for covariates, there may be other factors that could have influenced the estimates. However, for the analysis of serious adverse events, we evaluated all diagnoses set during hospital stay both before and after the first dose of vaccine, thereby increasing the chance of detecting selection bias that could interfere with the results. In addition, the sensitivity analysis wherein influenza was used as a negative control outcome supported the lack of important confounding.

Moreover, because COVID-19 hospitalisation in this population was very rare, it was not possible to estimate the VE of a first booster dose in this cohort with any form of precision. Strengths of this study include that all the registers used to obtain the data used in the present study have nationwide coverage with virtually zero loss to follow-up. Finally, the study cohort was based on the total population of Swedish adolescents, including 0.8 million individuals aged 11-19 years of age, which increases the possibility to generalize the findings to other countries with similar population structures.

## Conclusion

In summary, COVID-19 mRNA vaccination was not associated with an increased risk for hospitalisation of any cause in adolescents, suggesting that they are safe to use. While vaccination was associated with a reduced risk of COVID-19 hospitalisation during the omicron era, the absolute risk among general adolescents was extremely low and similar to that of influenza. In contrast, in a large proportion of adolescents hospitalised due to COVID-19, certain risk factors were present, and the effectiveness of vaccination in these individuals was similar to in the total cohort. These results indicate that certain vulnerable subgroups of adolescents, rather than adolescents in general, should be prioritized for vaccination.

## Data Availability

The data files used for the present study are publicly unavailable according to regulations under Swedish law. However, all data used for the present study can be applied for from the National Board of Health and Welfare, Statistics Sweden, and the Public Health Agency of Sweden.

## Contributors

PN conceived and designed the study. PN acquired the data. PN performed the statistical analyses. PN and MB accessed and verified the underlying data. All authors interpreted the data. PN and MB drafted the manuscript. All authors critically revised the manuscript for intellectual content. PN and AN supervised the work. All authors gave final approval of the version to be published. All authors had full access to all the data and had final responsibility for the decision to submit for publication

