## Supplementary table 1 for "Safety and effectiveness of COVID-19 mRNA vaccination and risk factors for hospitalisation caused by the omicron variant in 0.8 million adolescents: A nationwide cohort study in Sweden"

### Contents

**Supplementary Table 1.** ........................................ **2**

**Supplementary Table 1. Type of SARS-CoV-2 genotypes based on whole genome sequencing in Sweden during the follow-up period of the present study (week 1-22, 2022). Data publicly available at the Public Health Agency of Sweden (https://www.folkhalsomyndigheten.se/smittskydd-beredskap/utbrott/aktuella-utbrott/covid-19/statistik-och-analyser/sars-cov-2-virusvarianter-av-sarskild-betydelse/)**

| | Type of SARS-CoV-2 variant | | | | | | | |
|---|---|---|---|---|---|---|---|---|
| Week number , 2022 | Delta (B.1.617.2) Number of cases | Omicron (B.1.1.529) Number of cases | Omicron (BA.1) Number of cases | Omicron (BA.2) Number of cases | Omicron (BA.4) Number of cases | Omicron (BA.5) Number of cases | Total number of whole genome sequenced | Proportion whole genome sequenced of all confirmed cases (%) |
| 1 | 365 | 233 | 3 158 | 474 | | | 4 353 | 3 |
| 2 | 173 | 320 | 3 468 | 905 | | | 4 999 | 3 |
| 3 | 54 | 323 | 2 827 | 1 458 | | | 4 876 | 2 |
| 4 | 21 | 132 | 2 447 | 2 061 | | | 4 876 | 2 |
| 5 | 5 | 229 | 2 223 | 3 237 | | | 5 942 | 3 |
| 6 | 1 | 258 | 1 328 | 3 113 | | | 4 864 | 8 |
| 7 | 0 | 238 | 865 | 2 612 | | | 3 916 | 18 |
| 8 | 0 | 168 | 563 | 2 908 | | | 3 758 | 21 |
| 9 | 0 | 105 | 369 | 2 972 | | | 3 536 | 28 |
| 10 | 0 | 136 | 215 | 2 899 | | | 3 331 | 33 |
| 11 | 0 | 111 | 116 | 2 405 | | | 2 684 | 32 |
| 12 | 0 | 132 | 45 | 2 061 | | | 2 248 | 32 |
| 13 | 0 | 139 | 21 | 1 548 | | | 1 723 | 35 |
| 14 | 0 | 115 | 16 | 1 260 | | | 1 416 | 35 |
| 15 | 0 | 16 | 11 | 1 077 | 0 | 5 | 1 159 | 39 |
| 16 | 0 | 1 | 4 | 917 | 0 | 3 | 954 | 40 |
| 17 | 0 | 1 | 2 | 727 | 2 | 1 | 741 | 36 |
| 18 | 0 | 0 | 4 | 677 | 5 | 7 | 702 | 41 |
| 19 | 0 | 0 | 2 | 742 | 11 | 11 | 781 | 54 |
| 20 | 0 | 1 | 1 | 658 | 21 | 26 | 727 | 53 |
| 21 | 0 | 1 | 0 | 564 | 27 | 58 | 659 | 56 |
| 22 | 0 | 4 | 0 | 518 | 30 | 118 | 675 | 55 |
